# Pilot implementation of short message service for randomisation in a multisite pragmatic factorial clinical trial in Kenya

**DOI:** 10.1101/2023.12.06.23299618

**Authors:** Mercy Chepkirui, Dennis Kimego, Charles Nzioki, Elizabeth Jowi, Charles Opondo, Ambrose Agweyu

**Author notes:** Author contributions: CO conceived the project, and MC and DK developed the SMS platform. AA and CO supervised the development of the application, The first draft of the manuscript was prepared by MC, and further developed by AA and CO. CN and EL provided clinical trial site supervision. All authors critically reviewed the paper before submission.

## Abstract

The traditional use of sealed envelopes for randomisation is susceptible to manipulation and the risk of damage to envelopes during shipping and at storage. Additionally, the filling and sealing envelopes is, tedious, time-consuming, and error prone. Other randomisation alternatives such as web-based methods are preferred. However, they are expensive and unsuitable in settings with poor internet infrastructure. Mobile phone-based randomisation using Short Message Service (SMS) potentially offers a low-cost and reliable alternative.

We developed an SMS-based method for random allocation of treatments. Plain text messaging or an Android app were used to formulate text messages using a fixed syntax consisting of participant unique identifier, trial site, stratum, and the trial name as input parameters. The system verified the input parameters and obtained an allocation from the database before returning a response to the sender. The text response contained the details of the treatment allocation. The study was done in two sites of a multi-site 3×2 factorial clinical trial in Kenya involving two interventions with up to nine possible allocations. We evaluated the accuracy of treatment allocations against the master randomisation list for each randomisation SMS processed, and SMS latency in seconds. A post-implementation survey was conducted to evaluate user feedback.

A total of 218 participants were randomised between 7th February 2022 and 11th April 2022, out of which 179 were randomised to only one arm while 39 were randomised to both treatment arms. Allocation accuracy was 100%. Median latency was 22 seconds with the fastest message processed in 10 seconds and the slowest (non-network delayed) message processed in 2129 seconds. Four users completed a post-implementation survey. The pilot study demonstrated that SMS randomisation to be easy, user-friendly, fast, and accurate and a feasible alternative randomisation technique.

**Author Summary:** While conducting a randomized clinical trial with a sample size of more than 4000 participants, in routine care settings in Kenya, we encountered a challenge with the approach to randomisation. In our study protocol we settled on the use of sealed opaque envelopes for allocation concealment. This approach, given our large sample size and nine possible treatment allocations, necessitated the preparation of an overwhelming (4000×9) envelope stratified across 12study sites. This manual process was tedious, time consuming and error prone, with issues arising such as mislabelling, empty envelopes, and some being damaged in storage or transit.

Recognizing these challenges, our team was prompted to innovate an alternative digital solution, with the ultimate aim of establishing a proof of concept that could support future clinical trials in routine care settings. Given the high mobile penetration in Kenya, we sought to leverage SMS-based mobile communication as a means of determining treatment allocation for study participants. We developed a platform capable of accommodating both Android and feature phones using open-source tools. Our findings indicate that SMS is a fast, user-friendly, and low-cost method, presenting a viable solution that could potentially revolutionize randomisation in clinical studies. However, it is important to note that our testing of this method was limited to only two study sites. Despite this, our study lays the groundwork for digital randomisation and hopefully inspire future advancements in health research.

## Introduction

Randomisation of participants in clinical trials has become the standard method of experimental control aimed at reducing selection bias and eliminating confounding from known and unknown factors(1). The process of randomisation generally involves two steps: (i) Generating an unpredictable sequence of random assignments, and (ii) Implementing the sequence in a way that conceals the treatment assigned to potential study participants until eligibility is determined(2,3). Failure to achieve proper randomisation and allocation concealment may result in biased estimates of treatment effects and potential loss of integrity of trial results(4).

Traditionally the use of sequentially numbered, opaque, sealed envelopes has been regarded an acceptable method for concealing allocation of interventions in trials. However, this method is now falling out of favour due to vulnerability to manipulation(5). Furthermore, sealed envelopes are susceptible to damage during shipping and storage. The process of filling and sealing envelopes is also a time-consuming manual process which is prone to human error, particularly in large complex studies.

In response to the limitations associated with sealed envelopes and recognising inadequate methodological approaches in controlled trials, there is a growing inclination towards the adoption of centrally administered web-based or telephone-based randomisation in large studies. However, implementing these methods is challenging in settings with inadequate communication infrastructure(6) and unreliable internet connectivity.

An alternative approach to randomisation, which is low in cost, auditable, and particularly suited for Low- and Middle-Income Countries where access to mobile phone technology has rapidly expanded(7,8), involves the use of mobile phone-based Short Messaging Service (SMS). SMS is a method of communication that transmits text messages up to 160 characters in length, between mobile devices or from a computer to a mobile device. Kenya is reported to have 98% mobile penetration amongst adults(9).

Bulk messaging enables the synchronous delivery of SMS text messages to a vast number of recipients minimizing delays and overlapping requests. In clinical trials, text messaging has proven effective in reducing missed appointments(10) and has served as a cost-effective intervention for managing patients with chronic illnesses(11–14).

We developed an SMS-based method for random allocation of treatments and subsequently undertook a pilot study comparing an SMS-based randomisation platform versus the conventional approach using sealed opaque envelopes. The study was conducted in parallel with a 3×2 factorial pragmatic randomised controlled trial of alternative treatments for severe pneumonia among children aged 2-59 months(15).

Our aims were to evaluate the feasibility and accuracy of randomisation using text messaging by estimating the response time of SMS delivery for randomisation requests, assessing the user experience for envelope randomisation and SMS randomisation approaches, correct treatment allocation, and determining allocation sequence concordance for envelope randomisation and SMS randomisation.

## Results

Between February 2022 and May 2022, 218 participants were successfully randomised in the two participating clinical trial sites using the SMS approach. One seventy-nine (82.1% - antibiotic arm) participants were randomized in one step and 39 (17.9% - antibiotic & supportive care arm) were randomized in two steps.

In the testing and pilot phases of the study, we logged a total of 580 SMSes, which we categorized as shown in Table 1. We noted various types of SMS requests. For 151 (26%) that fell under the *’invalid non-authorized request*’ category, messages consisted of syntax completely unrelated to randomisation, often missing the keyword ‘*randomize*’. The system reported 22 (4%) requests attempting to randomize participants who were already allocated treatments. One SMS reported under the ‘*exhausted sequence’* category was a test case scenario where the allocation sequence was no longer available for randomisation. Two unregistered users made attempts to randomise participants, while 402 (69%) requests had valid syntaxes that were processed, and an allocation treatment was delivered to the user as a response.

**Table 1:**
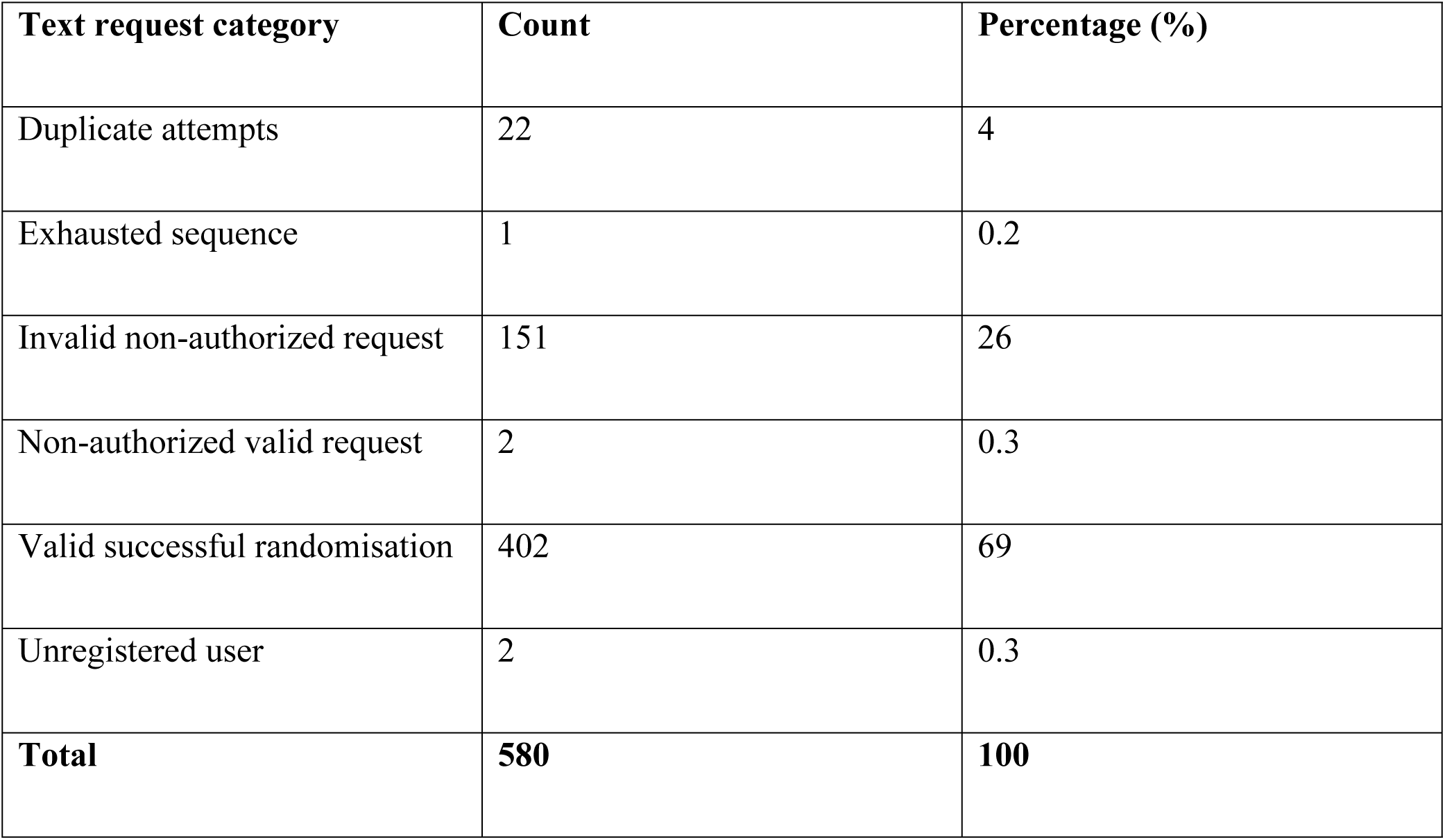
Total SMSes processed during the testing and piloting phases of the study.

| Text request category | Count | Percentage (%) |
| --- | --- | --- |
| Duplicate attempts | 22 | 4 |
| Exhausted sequence | 1 | 0.2 |
| Invalid non-authorized request | 151 | 26 |
| Non-authorized valid request | 2 | 0.3 |
| Valid successful randomisation | 402 | 69 |
| Unregistered user | 2 | 0.3 |
| **Total** | **580** | **100** |

The SMS latency for the valid successful randomisation processed requests are as shown in Table 2. The median latency was 22 seconds, with the fastest processed SMS taking just 10 seconds (IQR 29.75 seconds). *We observed one delayed response, which was eventually delivered 35 minutes later.* It stands out as an outlier as majority of the SMSes were processed under 100 seconds.

**Table 2:** SMS latency IQR table for valid successful randomisation requests.

| **Latency range** | ***Total SMSes*** | ***Min.*** | ***1st Qu.*** | ***Median*** | ***Mean*** | ***3rd Qu*** | ***Max.*** |
| --- | --- | --- | --- | --- | --- | --- | --- |
| **Less than 100 secs** | *393(98%)* | *10.00* | *16.00* | *21.00* | *31.65* | *44.00* | *96.00* |
| **All** | *402* | *10.00* | *16.00* | *22.00* | *46.05* | *45.75* | *2129.00* |

Allocation accuracy was 100% when compared to the allocation sequence.

Four clinicians completed a post-pilot survey. From the responses it took a clinician less than two minutes to compose a randomisation text. Two exclusively used the mobile app for randomisation, while two utilized both feature phones and the app. Generally, the clinicians reported that they found SMS randomisation easy to grasp and use. However, opinions on preference were split; two clinicians favoured envelopes, while two preferred text messaging. The Android application was notably preferred over manual texting for composing the randomisation texts. A recurring challenge was forgetfulness in using text randomisation.

## Discussion

Our research provides unique insights as, to the best of our knowledge, it is the first study investigating a Mobile SMS randomisation approach in a low-income setting within a complex randomized controlled clinical trial. Envelope randomisation, a manual and traditional method, is reliant on the integrity of filling, sealing, transporting and storage of envelopes, highlighting the need for digital alternatives such as SMS. In our pilot study, we found SMS randomisation to be user-friendly, efficient, fast, and accurate. It also addressed significant challenges associated with envelope randomisation, such as the time-consuming process of preparing envelopes and uncertainties related to envelopes being unsealed or damaged. Additionally, it eliminates the needs for paper, printing, and shipping.

Given the high mobile penetration and widespread use of SMS messaging (16) with a the low cost of SMS at $0.0078 per unit, the potential of SMS randomisation is evident. SMS processing through a local network recorded a turnaround time of 22 seconds. This highlights the approach’s practical potential in pragmatic trials, ensuring that there are no delays in service delivery within a busy public routine care hospital setting during SMS randomisation. The introduction of the mobile application that automated SMS formulation made randomisation more efficient, and no internet connectivity was needed. Despite the predominance of feature phones in the Kenyan Market(17), our solution demonstrates versatility, proving that mobile applications and feature phones can be seamlessly integrated and used interchangeably for SMS randomisation. This ensures broad accessibility and efficiency across different device types.

### Digital randomisation techniques

As randomisation is a key determinant of the effectiveness of a clinical trial, trialists need to embrace improved and innovative methodologies that include the use of technology where applicable. Trialists are increasingly exploring digital options for conducting randomised controlled trials to mitigate challenges affecting recruitment in clinical trials(18–20). Digitization stands out in ensuring correct and accurate treatment allocation. This not only helps in maintaining the integrity of the clinical trial but also plays a crucial role in minimizing and scrutinizing potential biases in trial outcomes, thereby enhancing the credibility of the results. Clinical trial monitoring becomes more efficient as trial progress can be readily traced in real-time through a randomisation dashboard integrated into the trial data collection process. This feature is particularly beneficial for adaptive clinical trial designs, where the ability to make data-driven decisions in real-time is paramount(21).

### Clinical trials in low-resource settings

Mobile-based randomisation can solve a number of clinical trial challenges inherent in low-resource settings such as financial constraints, operational barriers such as remote locations of study sites, and limited human capital (22). Setting up clinical trials with complex designs can be prohibitively expensive in such settings (23). This calls for effective methods of conducting trials that deliver credible results while minimizing cost. Our approach, developed using open-source tools, serves a testament to the feasibility of digitizing clinical trial methods in low resource settings. Representing marginalised populations in health research and innovation is crucial for addressing the significant disease burden in low-income countries in a fair and equitable manner (22,24–26). There is an urgent need for investment in solutions that will increased the number of clinical trials conducted in low-income countries (27). As our approach only targets two components of clinical trials – randomisation and trial monitoring – additional research is required to pilot other low-cost tools that could improve the quality of clinical trials in similar contexts.

### Limitations and recommendations

We acknowledge various limitations to our study. The pilot was done in a restricted context with limited number of users and trial sites. Users were clinicians already involved in the larger randomized controlled trial, which could have influenced their feedback and experience. There may be learning curve or initial hesitations for naïve. Clinicians admitting to often forgetting to send the text request after opening the envelope also highlights a behavioural aspect that could be addressed in future implementations to ensure consistent use of the system. The application logic was informed by a simple randomisation technique and tested in an urban setting in Kenya setting with local SMS service providers. This paper does not extend the discussion to implementation in other countries or in rural Kenya where network connectivity may be unstable. Nonetheless, our findings are promising and recommend conducting pilots in various settings, clinical trial designs, and geographical locations. Future iterations of the SMS-based system could introduce enhancements that optimize reliability and guarantee integrity.

### Conclusions

The promising results from our pilot indicate that there is potential for wider implementation in large-scale clinical trials. The observed improvements in efficiency, high accuracy, and user acceptance point to the viability of SMS randomisation in clinical research in both low and high resource settings. We used open-source tools for the development and testing of the SMS platform, ensuring accessibility for further development and improvements. This lessons from this trial serve as a reference point for future low-cost technology-driven innovations to expand the reach and quality of clinical trials globally.

## Materials and methods

We conducted a prospective two-arm pilot study nested within an actively recruiting randomised controlled trial. This study was conducted in two phases. The development phase (Phase 1) involved the design specification of the SMS platform, and initial testing in web-based, text messaging and Android applications. The implementation phase (Phase 2) involved deployment of the application at two public hospitals in Kenya: Machakos Level 5, and Mama Lucy Kibaki Hospitals, selected purposively from a pool of 12 clinical trial sites due to their high participant recruitment rates.

### Phase 1: Development phase

We designed and developed a three-tier SMS-based randomisation system consisting of data, application, and presentation interfaces (Figure 1). The requirements of the application were derived from standard operating procedures for randomisation in the larger clinical trial. Therefore, the logic was structured to accommodate a multi-step factorial randomisation design involving two interventions with up to nine possible allocations (Figure 2).

**Figure 1:**
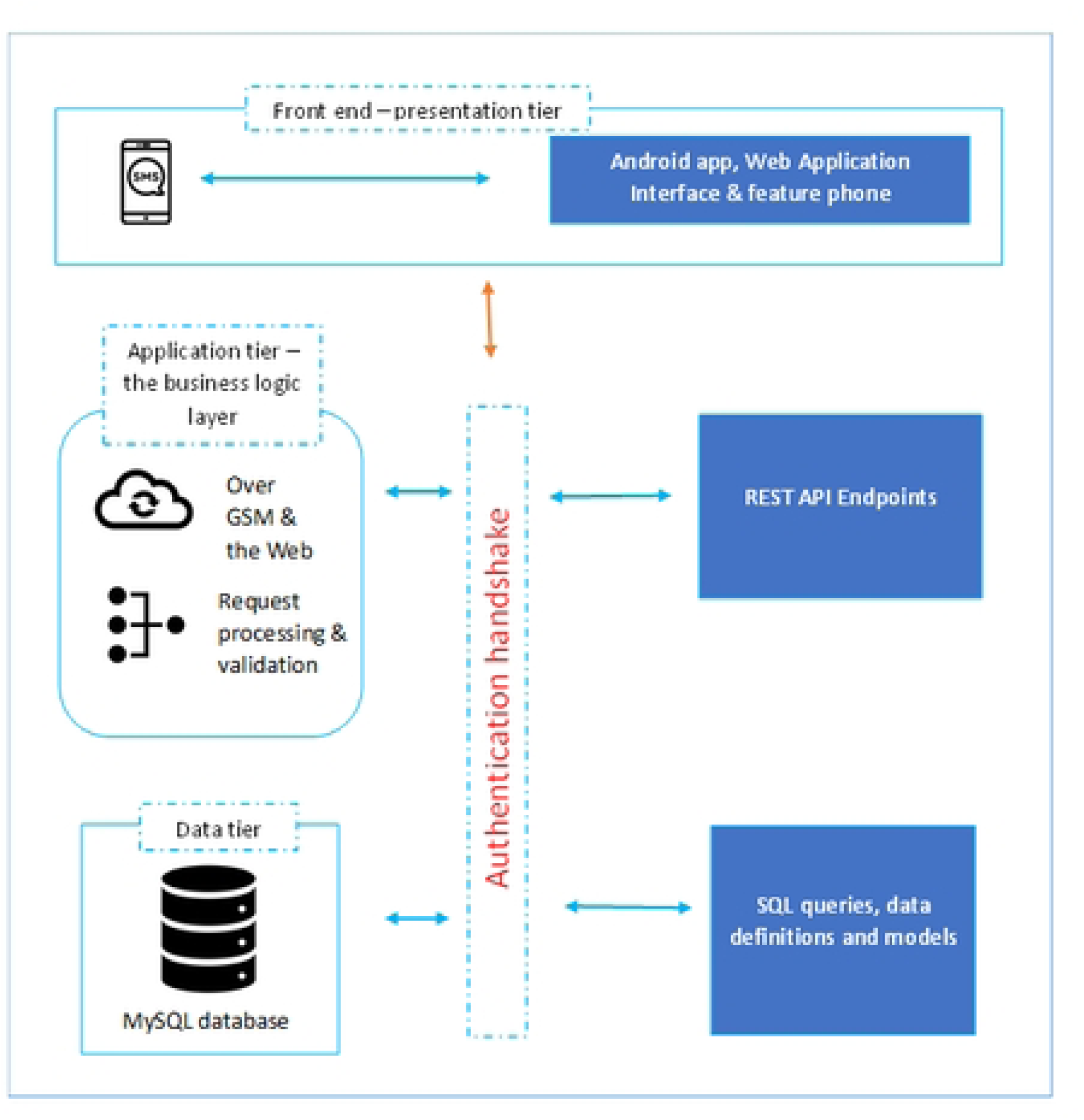
SMS platform design framework. The data tier stored all the system data, the business logic tier processed all the system transactions, and the presentation tier was the point of interaction between the user and the system.

**Figure 2:**
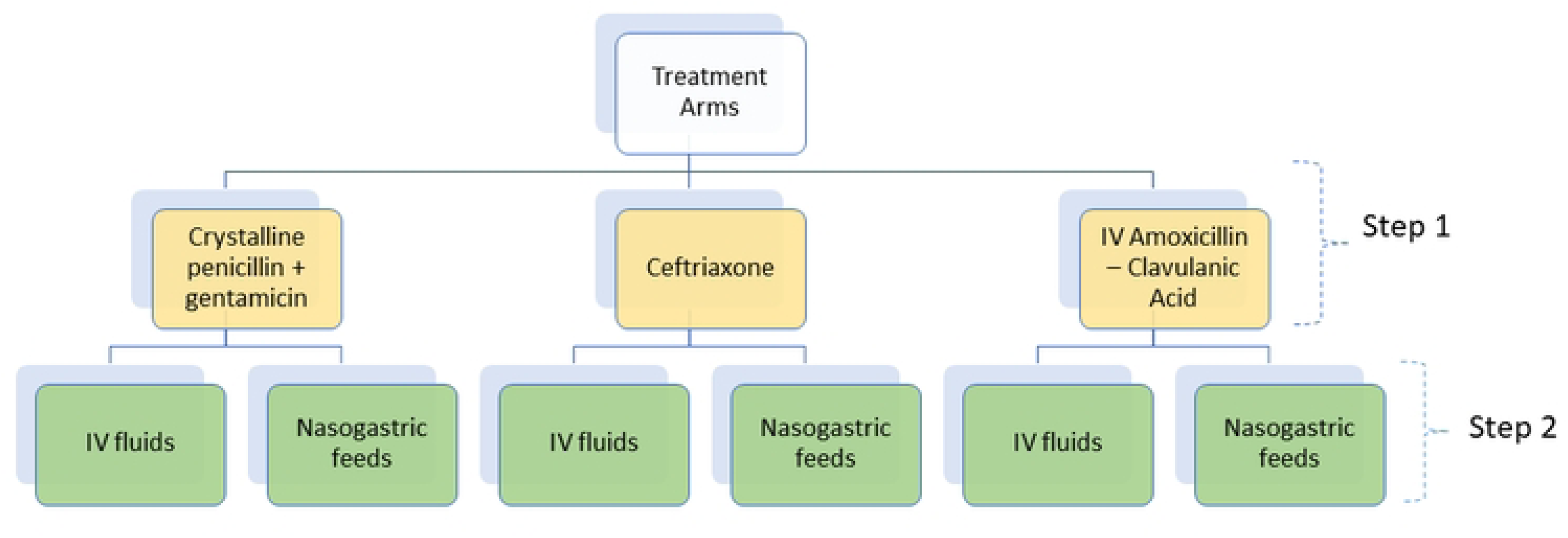
Factorial allocation of treatments. Three antibiotic treatment arms (crystalline penicillin & gentamicin, ceftriaxone, and Intravenous (IV) amoxicillin-clavulanic acid) and two supportive care treatment arms (Nasogastric feeds and IV fluids).

Each message consisted of a predefined ordered syntax comprising the participant unique identifier, trial site, stratum, and trial name. Detailed descriptions of the syntax, message scenarios, and expected responses are provided in Table 3 and Table 4.

**Table 3:** SMS formulation syntax of a randomisation request.

| Type of message | Input parameters formats | Example text |
| --- | --- | --- |
| A request to randomise a participant in each stratum at a trial site in a multisite clinical trial. | Randomise [IPNO] to [study name] [sitename] [stratum name] | *{Randomise RL567 to SEARCH MKSRT supportive}*<br>RL567 – IPNO<sup>1</sup><br>SEARCH – Study name<br>MKSRT – Site name<br>Supportive – stratum |
| A request to randomise a participant in each site (not stratified) in a multisite clinical trial | Randomise [IPNO] to [Study name] [sitename] | *{Randomise KL789 to SEARCH MMLY}*<br>KL789 – IPNO<br>SEARCH – study name<br>MMLY – site name |

<sup>1</sup> The unique patient identifier primarily assigned to a patient at the public hospital.

**Table 4:** Various SMS formulation request scenarios and their respective expected responses.

| **Event scenario** | **Expected response** |
| --- | --- |
| **Non-registered user** | *The number [phone number] does not belong to an active user who is authorised to randomise participants study at [site name]. Contact [administrator contact] for more details.* |
| **Exhausted allocation list** | *Random allocations to the [study name] study is no longer available. Please contact the study co-ordination centre [Phone number].* |
| **Invalid message format** | *Incorrect message format; use: randomise [IPNO] to [studyID] [siteID] or: randomise [IPNO] to [studyID] [siteID] [phoneNO] without the straight brackets. You may also add your phone number at the end of the message if using an authorised phone that does not belong to you.* |
| **An attempt to randomise a participant twice** | *The participant with the [IPNO] is already allocated [allocation] by [username] at [timestamp].* |
| **An attempt to deactivate non-existing user** | *Deactivation failed; there is no record with the phone number [phone number] in the list of users.* |
| **Successful user deactivation** | *The user with the phone number [phone number], is now an inactive user.* |
| **Successful user registration** | *[Username] phone number [phone number], has been added to the list of users authorised to randomise participants to the [study name] study at the [site name] site.* |
| **Delete user** | *The phone number [phone number], has been removed from the list of users.* |
| **An attempt to add user twice** | *[username], phone number [phone number], already exists in the list of users.* |
| **Successful randomisation** | *Participant [IPNO] has been randomised to [allocation] in the [study name] study. The unique number for the participant is [participant randomisation ID]. Randomised by [trial staff name] on [timestamp].* |

The application tier verified the input parameters received from the mobile network operator through SMS or HTTP via a Representational State Transfer Application Programming Interface (REST API), obtaining an allocation from the data tier stored on a local database. It then returned a response to the sender through an SMS. The text response contained details of the treatment allocation, participant identifier, and identity of the study staff undertaking randomisation. The system was designed to identify duplicate randomisation attempts using unique patient identifiers (IPNO).

The randomisation application logged all the SMS processed (Table 5). These included invalid text messages, duplicated attempts to randomise, non-authorized request from users not registered and successfully processed valid randomisation requests. Valid randomisation and allocations were logged and captured in the administrative dashboard for review during trial monitoring. This log captured allocations for the two clinical trial arms – *antibiotic care and supportive care*. Antibiotic treatment allocation was the first step of randomisation consisting of three antibiotic regimens: crystalline penicillin and gentamicin, ceftriaxone, and intravenous (IV) amoxicillin-clavulanic acid. Supportive care arm was allocated as the second randomisation step consisting of two treatments: Nasogastric (NG) feeds and IV fluids (Figure 2).

**Table 5:** SMS requests categories.

| No. | Category | Definition |
| --- | --- | --- |
| 1 | Duplicate attempts | A SMS request that attempted to randomize a participant who had already been allocated a treatment in any arm. |
| 2 | Exhausted sequence | An attempt to randomize when an allocation sequence had been exhausted or fully utilized. |
| 3 | Invalid non-authorized request | A non-authorized user attempted to randomize a participant by sending a non-structured SMS. |
| 4 | Non-authorized valid request | A non-authorized user attempted to randomize a participant by sending a correctly structured SMS. |
| 5 | Valid successful randomisation | A randomisation attempt that was processed and a treatment allocation was sent out as a response to the user. |
| 6 | Unregistered user | A user who had not been registered attempts to randomize a participant. |

The administrative dashboard was a central hub for monitoring all transactions and randomisation logs. It stored the randomisation sequence, which was uploaded, during set up allowed administrators to supervise trial randomisation, manage users, and track recruitment across different sites and strata. A feature phone and the mobile application served as randomisation access points.

The mobile application was developed in Java for Android, and the web-based platform was developed using the PHP Laravel framework. The platform integrated with an SMS Application Programming Interface (API) from a local premium rate service provider (PRSP). One local mobile network operator was chosen for piloting due to cost related estimations. The source code for the SMS dashboard and the mobile application of this project is archived on GitHub (28,29). The web-based administrative dashboard is locally hosted on the KEMRI-Wellcome Trust servers following data management procedures outlined in the study protocol. At the time of development of this manuscript, the mobile application had not yet been published on the Google Play Store.

### Phase 2: Pilot SMS randomisation

Four clinical trial clinicians carried out the SMS randomisation pilot, with 2 clinicians stationed at each trial site. All users underwent training on how to use the SMS randomisation prior to piloting. The SMS platform was implemented in two modes: through text messaging on feature phones and smartphones using an Android mobile application. SMS randomisation was conducted alongside the traditional method of using envelopes. Each clinician was provided with a tablet computer with SIM card registered to the study. The custom Android application was installed on each of the tablets with each clinician having a separate account with a designated role to randomise participants. All system users were pre-registered in the database.

A study clinician would first screen patients for eligibility and then proceed to randomise them using sealed envelopes (the primary method) and finally repeat the process using text messaging. Randomisation requests were submitted in structured text format, either manually typed in the phone’s default text messaging application or formulated automatically by the Android application. Texts from the Android app included a phone number at the end, while manually typed texts did not. Randomisation marked the final step in recruitment before a treatment was allocated to a participant. Treatment allocation and administration was based on the envelope concealment method.

Patient care was always the priority, ensuring that the study procedures did not delay or interfere with treatment. There was no direct risk to participants from the procedures of this study. If technical issues arose, the clinicians were able to call the user support team at the KEMRI-Wellcome Trust Programme for help. Additionally, weekly review meetings were held to assess progress and address any emerging challenges. A post-implementation survey was used to evaluate user feedback. We built a user feedback into the mobile application, which only became active after the pilot implementation was completed. Each user of the randomisation module completed the questionnaire. The study covered the cost of the premium SMS subscription package, ensuring that users did not incur any additional charges.

### Data analysis

The SMS platform logged data for each SMS request made, capturing both the initiation time of the request and the time a response was delivered to the user. This enabled us to calculate a turnaround time, or SMS latency, in seconds for each processed message. We then analysed the data by computing the medians with interquartile ranges (IQRs) for turnaround time in seconds. To better describe the range of SMS requests made, we grouped all requests into 6 distinct categories with each SMS assigned to a group (Table 5). We only computed the SMS latency for valid randomisation requests (group 5 in Table 5). A valid request was defined as by the correct structured syntax with all the input parameters required for randomisation.

To determine the validity of an (In-Patient Number) (IPNO), we extracted all IPNOs in each SMS request and compared them against the IPNOs in the clinical trial database. We evaluated the accuracy of treatment allocations by comparing SMS request response for treatment allocation with the master randomisation list for each processed message. Survey responses from all the users were reviewed and summarized.

## Acknowledgments

This project would not have been possible without the kind of support and help from many individuals. We would like to extend our sincere gratitude to the following partner: SEARCH Clinical Trial Management Group, Mama Lucy Kibaki Hospital study clinicians, paediatric team, and data clerks, Machakos Level 5 Hospital study clinicians, paediatric team, and data clerks, and the KEMRI – Wellcome Trust Operations Department.

## Ethics approval and consent to participate

The Kenya Medical Research Institute (KEMRI) Scientific and Ethics Review Unit approved the collection of the deidentified data analysed in this study. The study clinicians consented to participate in the SMS randomisation pilot and to use their names and email addresses for SMS notification and verification purposes.

## Availability of data and materials

The data utilized in this work was generated from the SMS randomisation system. Further access to the data and additional system design materials can be sought through a request to KEMRI Wellcome Trust Research Programme’s Data Governance Committee through.

## Supporting information

**S1 Table. Total SMSes processed during the testing and piloting phases of the study.**

**S2 Table. SMS latency IQR table for valid successful randomisation requests.**

**S3 Table. SMS formulation syntax of a randomisation request.**

**S4 Table. Various SMS formulation request scenarios and their respective expected responses.**

**S5 Table. SMS requests categories.**

